# Unique Geographical Features of Out-of-Hospital Cardiac Arrest Patients within urban area: A Bayesian Spatial Analysis

**DOI:** 10.1101/2023.07.05.23292271

**Authors:** Atsushi Senda

**Affiliations:** Graduate School of Medical and Dental Sciences, Tokyo Medical and Dental University, Tokyo, Japan

**Keywords:** out-of-hospital cardiac arrest, Integrated Nested Laplace Approximation, Stochastic Partial Differential Equation, spatial modeling, urban, gaussian process, Bayesian, Japan, medical control

## Abstract

**Background:** The advantages of urban areas for patients with out-of-hospital cardiac arrest (OHCA), attributable to their extensive medical resources, are well recognized. However, whether a greater abundance of these resources directly improves patient outcomes is unclear. Moreover, it is important to clarify this because of the ongoing global trend of urbanization. Therefore, this study aimed to investigate this issue and shed light on the potential challenges specific to urban environments.

**Methods:** This retrospective observational study was conducted to evaluate the correlation between the geographical features of patients with shockable OHCA and neurological outcomes. Data of patients who were transported to Tokyo Medical and Dental University Hospital between June 1, 2016, and May 30, 2022, were extracted from electronic review board records. The Glasgow-Pittsburgh Cerebral Performance Category Scale was utilized to evaluate the neurological results. The study employed Bayesian spatial modeling and analyzed the results using the Integrated Nested Laplace Approximation and Stochastic Partial Differential Equation methods.

**Results:** Paradoxically, a region with the highest concentration of advanced medical facilities exhibited the poorest neurological outcomes. This area was characterized by an extended duration of on-site emergency medical service activity, which strongly correlated with a negative impact on patients’ neurological outcomes.

**Conclusions:** The abundance of healthcare resources in urban areas does not necessarily correlate with improved outcomes for patients with OHCA. A strategic approach to medical control that considers these factors can potentially enhance the outcomes of patients with OHCA in urban areas.

**Clinical Perspectives:** *What is new?:* In a retrospective geographical analysis of patients with shockable Out-of-Hospital Cardiac Arrest (OHCA), it was found that the region with the highest concentration of advanced medical facilities paradoxically exhibited the poorest neurological outcomes. Additionally, there was a noticeable extension in the duration of on-site emergency medical service activity in this area.

*What are the clinical implications?:* The abundance of healthcare resources in urban areas does not necessarily equate to improved outcomes for patients with OHCA. Therefore, a strategic approach to medical resource management should be considered to effectively utilize these resources and enhance the outcomes of patients in urban areas.

## Introduction

Despite advancements in emergency medicine, the survival rate and neurological outcomes for out-of-hospital cardiac arrest (OHCA) are suboptimal.^1,2^ The prognosis for OHCA is influenced by various factors, including age,^3^ sex,^4^ and comorbidities.^5^ Prompt action by bystanders and emergency medical services (EMS),^6–9^ coupled with comprehensive hospital treatment, can enhance neurological outcomes.^10,11^ Furthermore, spatial factors such as population density^12^ and rural–urban differences^6,13^ significantly affect the prognosis of OHCA. These findings highlight the advantages of urban environments, primarily due to the availability and accessibility of extensive medical resources.

There are trends of urbanization worldwide, and it is presumed that nearly 10 billion people are projected to live in urban areas by 2050.^14,15^ Tokyo, one of the world’s most densely populated urban areas,^16^ has over 250 active EMS units and more than 10 tertiary emergency hospitals, specializing in treating life-threatening conditions.^17^ This level of preparedness makes Tokyo a prime exemplar of urban readiness for OHCA incidents.

Previous studies have demonstrated the advantage of urban areas over rural areas.^6,13,18–20^ However, there is no previous study that evaluated the association between OHCA prognosis and geographical factors in urban settings, and it remains unclear whether the greatest abundance of these resources in city centers will lead to the best patient outcomes. Therefore, this study aimed to elucidate the correlation between the prognosis of OHCA and geographical factors in urban settings, a challenge that is poised to become a global concern.

## Methods

### Study design and setting

This retrospective observational study aimed to evaluate the correlation between geographical features and the prognosis of patients with OHCA exhibiting a shockable rhythm who were transported to Tokyo Medical and Dental University Hospital (TMDU) between June 1, 2016 and May 30, 2022. TMDU is a tertiary hospital located in the heart of Tokyo, in close proximity to the Imperial Palace, Tokyo Station, and central government offices. In Tokyo’s emergency system, hospitals are classified into three distinct levels based on the projected severity of the patient’s condition.^21^ TMDU, as a tertiary hospital, specializes in treating severe, life-threatening conditions, and as such, patients with OHCA are typically directed there.

This research followed the guidelines outlined in the 1964 Declaration of Helsinki and its subsequent revisions, and received approval from the Institutional Review Board of TMDU (#M2022-117). Given the retrospective design of the study and use of anonymized patient and hospital data, the requirement for informed consent from patients was waived.

### Study populations

This study enrolled patients with OHCA who exhibited shockable rhythm (ventricular tachycardia or ventricular fibrillation) on the arrival of the EMS team and were in a state of cardiopulmonary arrest on hospital arrival. The study excluded patients aged <18 years and those with existing “do not attempt to resuscitate” (DNAR) orders.

### Data collection

The data for this study were obtained retrospectively collected from the medical records of the patients: age, sex, Glasgow–Pittsburgh Cerebral Performance Category (CPC) scale score 30 days after hospital admission, EMS time-related details (time of EMS request, EMS arrival, on-site departure, and hospital arrival), and EMS geographical data (name of building, town, street, and address).

### Outcomes and definitions

The primary analysis involved the geographical assessment of neurological outcomes. Additional evaluations of the EMS on-scene response time (the duration from the EMS team’s arrival at the scene to departure) and transport time (the duration from departure from the scene to arrival at the emergency department) were performed. Neurological outcomes were assessed using the CPC scale. A score of CPC 1 signifies consciousness with normal function or only slight disability, CPC 2 represents consciousness with moderate disability, CPC 3 indicates consciousness with severe disability, CPC 4 denotes coma or vegetative state, and CPC 5 stands for death.^22^ For the analysis, patients were divided into those with CPC 1 or 2 (signifying good neurological outcomes) and those with CPC 3, 4, or 5 (indicating poor neurological outcomes).

### Statistical analysis

#### Bayesian spatial model

We assumed that there is a possibility of good neurological outcome *P*(*x*_i_) at each data point *x*_i_. By denoting the number of people *N*_i_at location *x*_i_, the number of people with good neurological outcomes (CPC 1 or 2) *Y*_i_ follows the distribution:

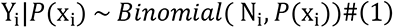

*P*(*x*_i_) at each location is modeled through the logit link function:

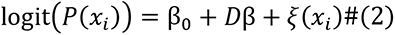

*β*_O_ denotes the intercept. Age and sex in each sample location were analyzed as covariates in the model and defined *β* = (β_age_, β_sex_) a vector of the covariate coefficients*. D* is a design matrix with rows corresponding to the covariate data for each sample location. Here, *ξ*(*x*) is a zero-mean Gaussian process with a Matérn covariance function^23^ as follows:

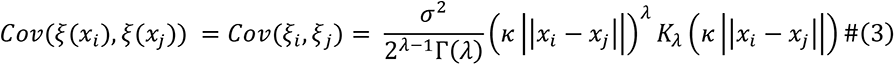

||*x*_i_ - *x*_j_||: Euclidean distance between locations *x_i_*, *x_j_*, *σ*^2^:marginal variance.

*K*_λ_ is the modified Bessel function of second kind, order *λ*(> 0): smoothness parameter, *κ*(> 0): scaling parameter related to the spatial correlation range.

#### Stochastic Partial Differential Equation approach

The Stochastic Partial Differential Equation (SPDE) approach enables approximation of a continuous process using discrete random process.^24^ SPDE can be written as

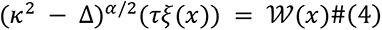

Where *s* ∈ ℝ^d^,Δ:Laplacian, *α*: smoothness parameter, *κ*: *scale parameter*, *τ*: variance,

𝒲(*s*): Gaussian spatial white noise process. This solution can be given by the Matérn covariance equation (3) and can be approximated using the finite element method, which can be represented by

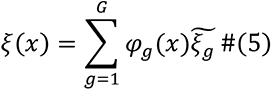

G: total number of vertices of triangulation, *φ*_g_: basis functions, 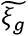 : zero-mean Gaussian- distributed weights.

#### Bayesian inference using Integrated Nested Laplace Approximation

The Bayesian inference was performed using Integrated Nested Laplace Approximation (INLA).^25^ The marginal posteriors of parameters *z* = (*β*_O_, *β*, *ξ*), and the hyper-parameters *θ*, can be represented by the following equation,

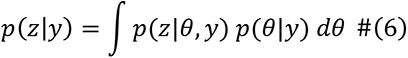

In the INLA approach, the integrand is approximated using the Laplace approximation, and numerical integration is performed. A detailed description is written elsewhere.^26^

#### Gaussian Process Regression between temporal information and neurological outcomes

Gaussian Process Regressions^27^ were conducted to elucidate the correlation between EMS on-scene response time and neurological outcomes and between transport time and neurological outcomes. The models were almost identical to equations (1) and (2); however, in this analysis, geographical information x_i_ was substituted with temporal information t_i_.

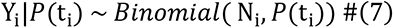

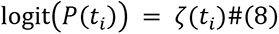

*ζ*(*t*) is a Gaussian process with Gaussian kernel:^23^

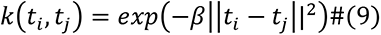

#### Software

The statistical analyses were conducted using R software (version 4.1.2; R Foundation for Statistical Computing, Vienna, Austria). Geospatial analyses were conducted using R-INLA (version 18.07.12). Google Maps Platform (https://mapsplatform.google.com/) was used to display the analyzed geographic information.

## Results

A total of 835 patients with OHCA were identified during the study period. Of these, 110 exhibited shockable rhythms on arrival of the EMS team. None of the patients was under the age of 18 years, and none had existing DNAR orders. Table 1 presents the demographic characteristics of the patients. The distribution of the 30-day CPC scores was as follows: CPC 1–18 (16.4%), CPC 2–16 (14.5%), CPC 3–14 (12.7%), CPC 4–12 (10.9%), and CPC 5–56 (50.9%).

**Table 1:**
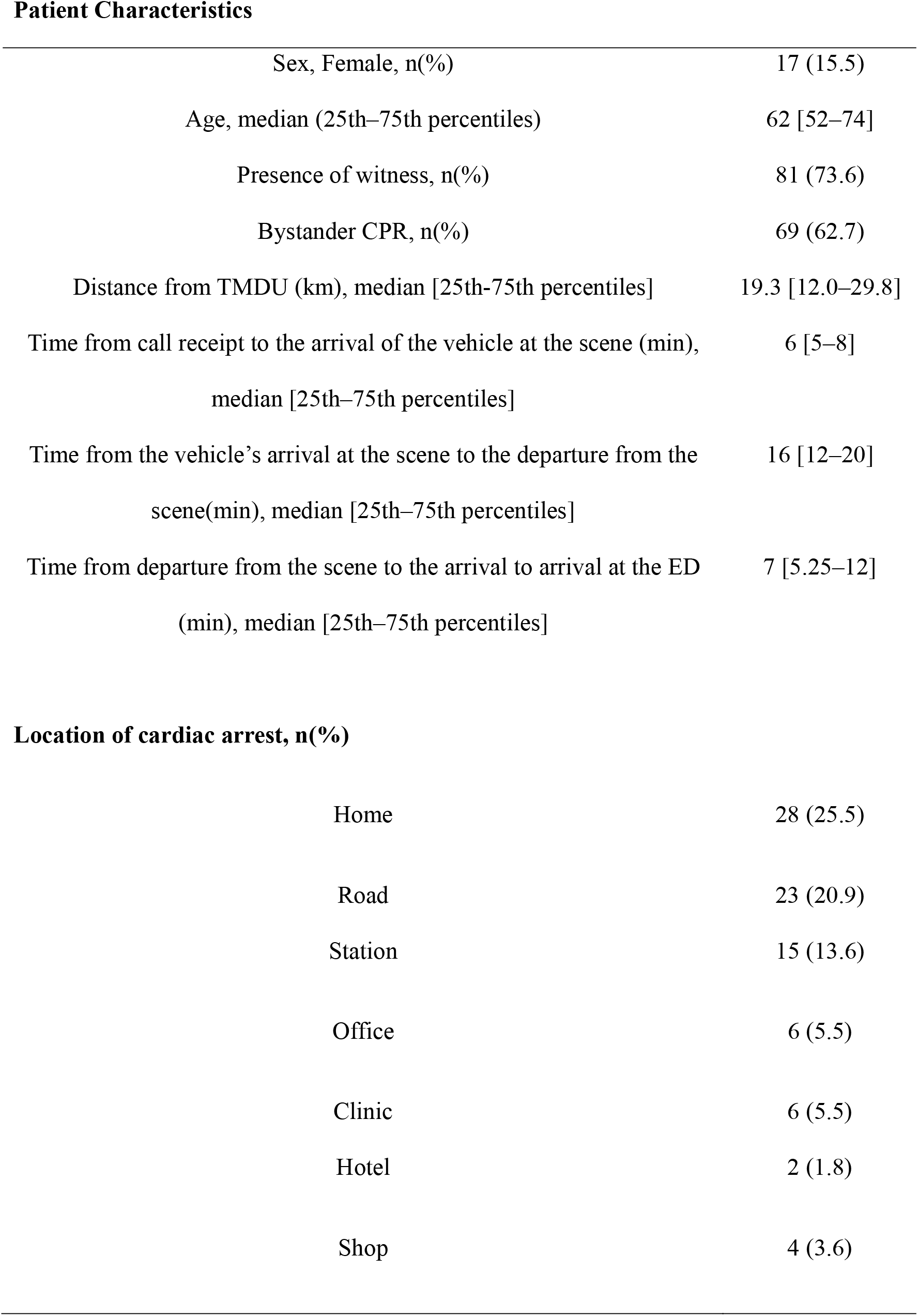
Patient demographics Abbreviations: CPR: cardiopulmonary resuscitation, ED: emergency department, TMDU: Tokyo Medical and Dental University

|  |  |
| --- | --- |
| Sex, Female, n(%) | 17 (15.5) |
| Age, median (25th–75th percentiles) | 62 [52–74] |
| Presence of witness, n(%) | 81 (73.6) |
| Bystander CPR, n(%) | 69 (62.7) |
| Distance from TMDU (km), median [25th–75th percentiles] | 19.3 [12.0–29.8] |
| Time from call receipt to the arrival of the vehicle at the scene (min),<br>median [25th–75th percentiles] | 6 [5–8] |
| Time from the vehicle's arrival at the scene to the departure from the<br>scene(min), median [25th–75th percentiles] | 16 [12–20] |
| Time from departure from the scene to the arrival to arrival at the ED<br>(min), median [25th–75th percentiles] | 7 [5.25–12] |
| **Location of cardiac arrest, n(%)** |  |
| Home | 28 (25.5) |
| Road | 23 (20.9) |
| Station | 15 (13.6) |
| Office | 6 (5.5) |
| Clinic | 6 (5.5) |
| Hotel | 2 (1.8) |
| Shop | 4 (3.6) |

### Neurological prognosis in relation to geographical features

The neurological outcomes of patients with OHCA and their correlation with geographical factors are presented in Figure 1 (figures without annotations are provided in Figure S1). Overall, there was a trend toward an improved neurological prognosis with increasing proximity to TMDU. However, it is noteworthy that despite being close to TMDU (2–8 km northeast of TMDU) and other tertiary hospitals, the region exhibited poor neurological outcomes (the area within the dotted line).

**Figure 1:**
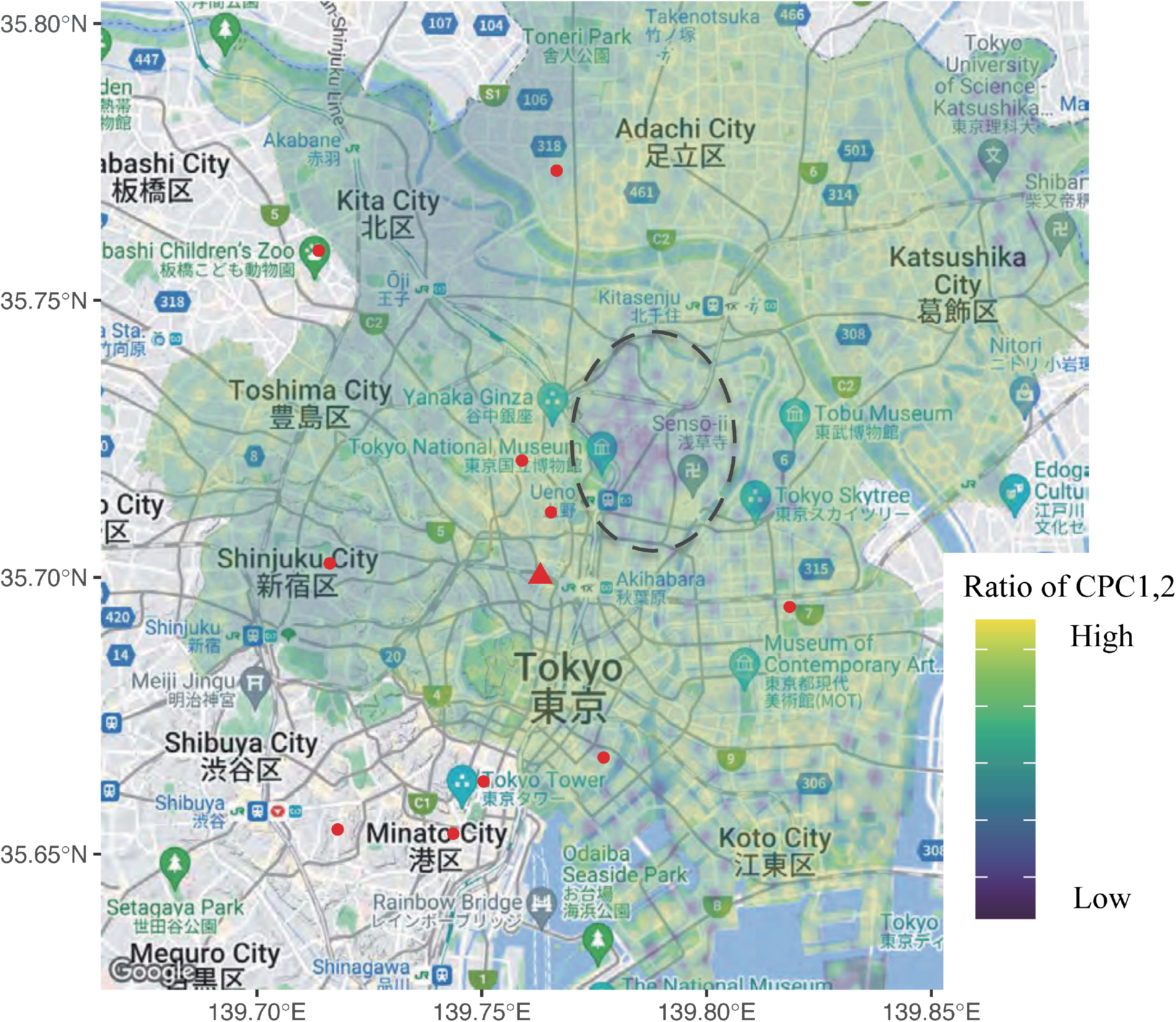
Neurological outcomes of patients with shockable rhythm out-of-hospital cardiac arrest in relation to geographical factors. The red circle denotes the location of tertiary medical centers, while the red triangle denotes the location of Tokyo Medical and Dental University. The area within the dotted line marks the region where neurological outcomes were poor despite its proximity to Tokyo Medical and Dental University. Abbreviations: CPC: Glasgow–Pittsburgh Cerebral Performance Category.

### Geographical differences in EMS on-scene response time and transport time

Geographical variations in the on-scene response time and transport time are depicted in Figure 2A and 2B, respectively. Figure 2B reveals a roughly positive correlation between the hospital’s distance and transport duration, with longer transport times associated with more distant locations. Figure 2A demonstrates the prolonged on-scene response times (median 18.0 min [25th–75th percentiles: 13.5–25.0 min]) within the area delineated by the dotted line in Figure 1.

**Figure 2:**
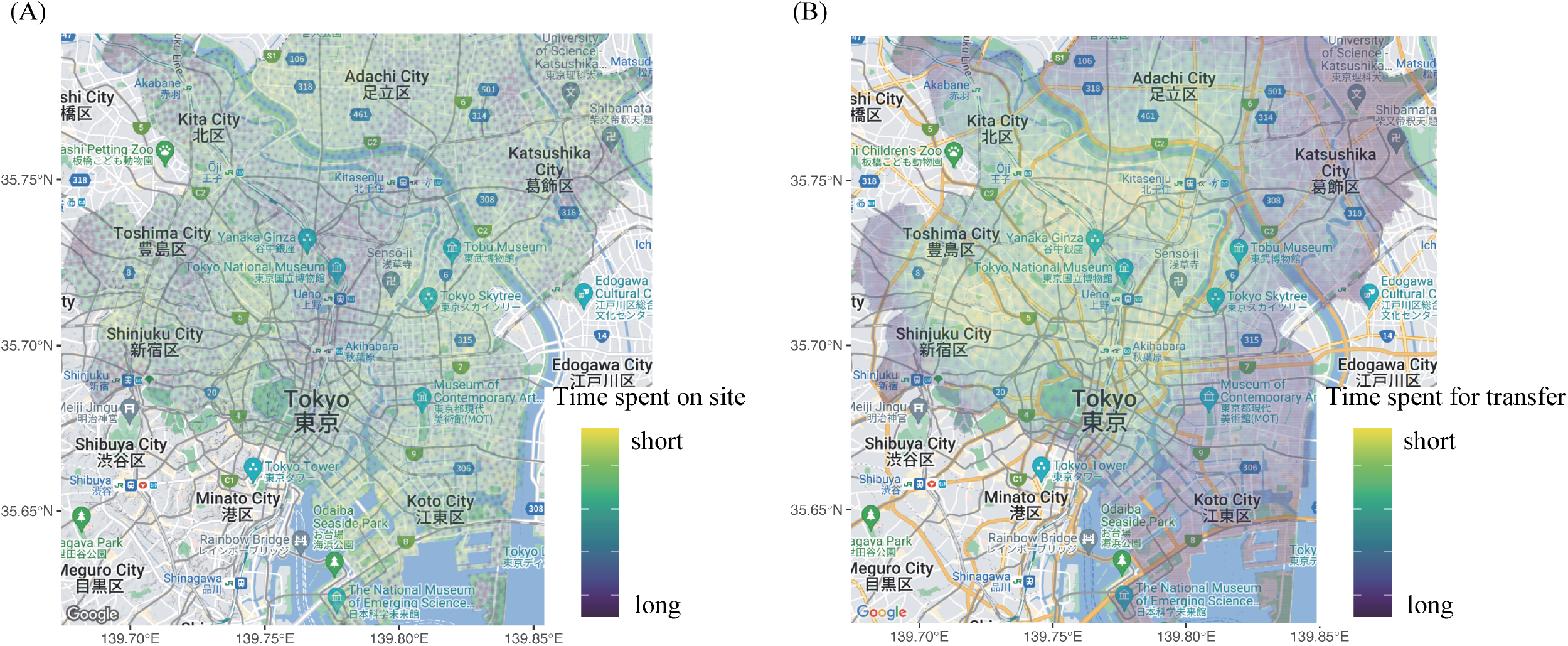
The on-scene response time (the time from the vehicle’s arrival at the scene to the departure from the scene) in relation to geographical factors (A). The transfer time (the duration from departure from the scene to arrival at the emergency department) in relation to geographical factors (B).

### Correlation between EMS on-scene response time/transport time and neurological outcomes

Figure 3A, B shows the correlations between on-scene response time and neurological prognosis and between transport time and neurological prognosis, respectively. Figure 3A illustrates that an increase in on-scene response time from 0 to 30 minutes results in a 50% decrease in patients with favorable neurological outcomes. Similarly, Figure 3B reveals that extending the transport time from 0 to 15 minutes leads to a 25% decrease in the percentage of patients with favorable outcomes. Both outcomes suggested that a longer duration was correlated with a worse prognosis. Notably, the on-scene response time appeared to have a more significant impact on neurological outcomes than the extended transport time.

**Figure 3:**
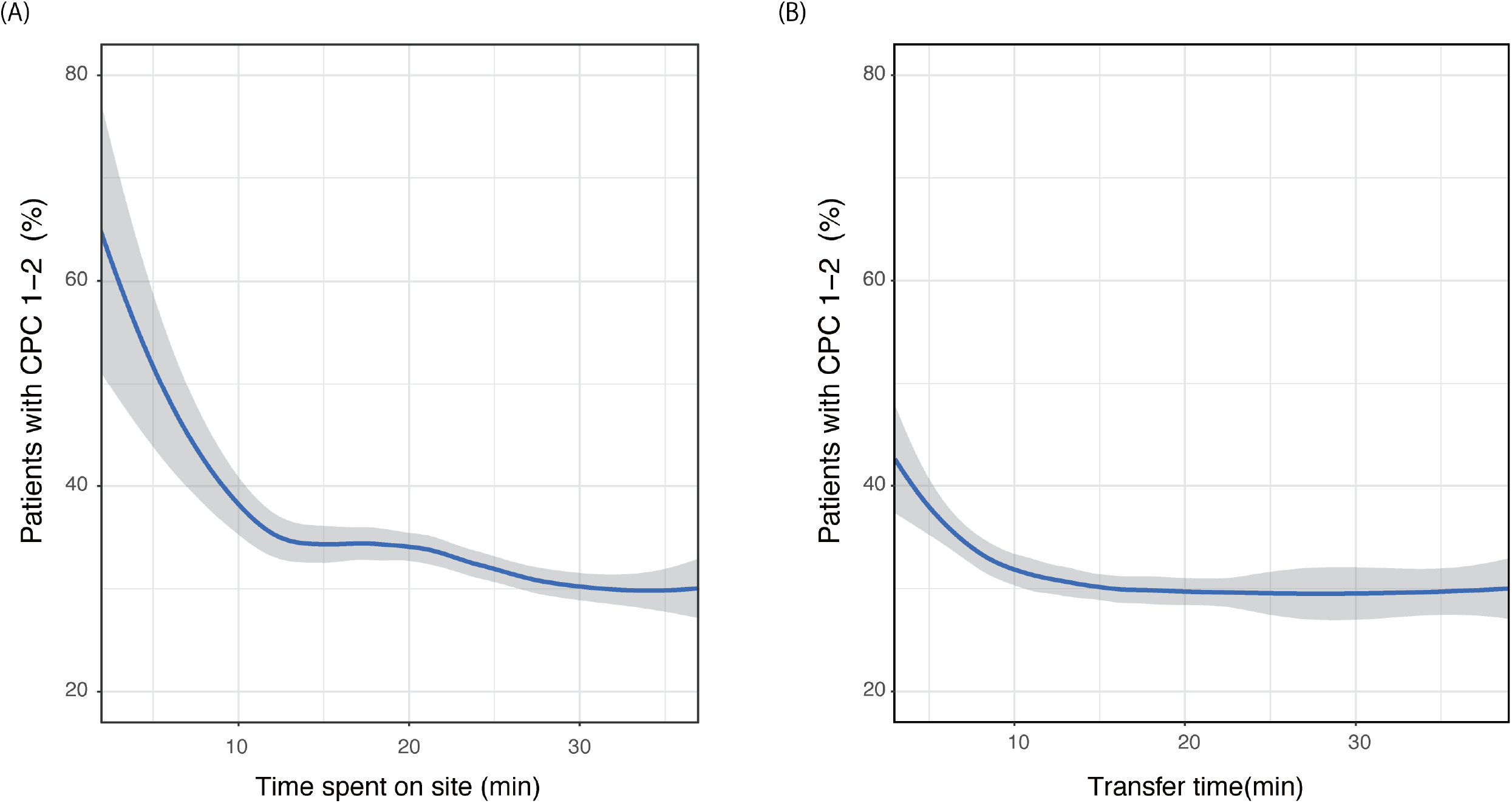
The relation between the on-scene response time (the duration from the EMS team’s arrival at the scene to departure) and neurological outcomes (A) and the relation between transport time (the duration from departure from the scene to arrival at the emergency department) and neurological outcomes (B). Abbreviations: CPC: Glasgow–Pittsburgh Cerebral Performance Category; EMS: emergency medical services

## Discussion

Previous studies have demonstrated a positive correlation between the degree of urbanization and the prognosis of patients with OHCA. These studies have generally compared urban areas with rural and/or suburban areas.^6,13,18–20^ However, to our knowledge, this is the first study that specifically investigates the regional variations within the urban areas themselves. We observed an area with a high concentration of advanced medical facilities that paradoxically demonstrated poor neurological outcomes. This area exhibited a longer duration of on-site EMS activity, which had a substantial negative impact on neurological outcomes.

Although most existing literature focuses on assessing mortality, the current study was designed to evaluate the neurological prognosis. This approach aims to mitigate perceived arbitrariness: a significant number of patients with poor neurological outcomes are faced with the decision to withdraw treatment.^28^ Moreover, we confined our study to OHCA cases involving patients who presented with shockable rhythms. This limitation was implemented because the initial rhythm poses a substantial confounder,^29^ which could influence the correlation between geographical features and patient prognosis even with adjustments.

In Tokyo’s emergency system, the selection of hospitals is made by sequentially contacting the facilities, starting from those nearest to the patient’s location, until a destination hospital is determined. Tertiary hospitals are densely situated in central Tokyo, with TMDU notably surrounded by these hospitals (Figure 1). Therefore, most OHCA cases in TMDU comprise patients who cannot be accommodated by other facilities. Another unique aspect of Tokyo’s emergency system is that EMS teams are rarely authorized to conduct basic life support. Certain EMS teams have the permission to place an adjunct airway, operate a semi-automated external defibrillator, and insert an intravenous line, but all these treatments are only authorized under the telephonic guidance of a doctor, with no other actions being permitted.^30,31^

Prior research suggests that extended transport time does not adversely affect survival rates or neurological outcomes.^32^ The consensus is that, despite prolonged transport times, the patient benefits from being transported to a high-level medical facility.^33^ The findings of the current study may seem to contradict this, but they can be better understood by considering two key points. First, this was a single-center investigation; hence, the type of medical institution did not act as a confounding variable between transport time and prognosis. Second, given the limitations of the resuscitation procedures that EMS teams in Japan can execute, extended transport times can lead to delays in the provision of advanced cardiovascular life support.

The prolongation of on-scene time negatively affects patient outcomes, which aligns with the findings of a previous study.^34^ Interestingly, extending the time on the scene appears to have a more detrimental impact on patient outcomes than the lengthening of transport time. This may be due to the fact that once the destination is determined, the hospital can prepare for the patient’s arrival, thereby making more efficient use of the transport time compared to the time spent on-scene.

This study has several limitations. First, although adjustments for confounders such as age and sex were made, the potential for unobserved and residual confounding factors could not be eliminated. Second, because this was a single-center study, we might not have captured geographical features that might only become apparent when collating data from multiple facilities. Third, our study was limited to patients with OHCA presenting with shockable rhythms. Therefore, our findings might not be generalizable to all patients with OHCA. Despite these limitations, our study is the first to focus on the demographics of patients with OHCA in urban medical systems. Poor neurological outcomes were apparent in areas with the most abundant medical systems. Based on the results of this study, it may be important to consider strategies that can effectively utilize abundant medical resources, such as pre-arranging the destination for patients with OHCA to avoid delays in transportation.

## Data Availability

The data are not publicly available due to privacy concerns regarding the research participants

## Acknowledgments

We would like to thank Editage (www.editage.com) for providing excellent assistance with the English language editing.

## Ethical approval statement

The study was conducted following the principles of the Declaration of Helsinki and was approved by the Institutional Review Board of Tokyo Medical and Dental University (#M2022-117; May 2023). The requirement for informed consent was waived because of the retrospective nature of the study and the use of anonymized data.

## Data availability

The data are not publicly available due to privacy concerns regarding the research participants

## Non-standard Abbreviations and Acronyms

CPC: Glasgow–Pittsburgh Cerebral Performance Category
DNAR: do not attempt to resuscitate
EMS: emergency medical services
INLA: Integrated Nested Laplace Approximation
OHCA: out-of-hospital cardiac arrest
SPDE: Stochastic Partial Differential Equation
TMDU: Tokyo Medical and Dental University Hospital

## Supplemental Material

Figure S1. The original of Figure 1 without annotations.

